# A Biallelic Variant of the RNA Exosome Gene *EXOSC4* Causes Translational Defects Associated with a Neurodevelopmental Disorder

**DOI:** 10.1101/2023.10.24.23297197

**Authors:** Milo B. Fasken, Sara W. Leung, Lauryn A. Cureton, Maha Al-Awadi, Adila Al-Kindy, Sohail Khoshnevis, Homa Ghalei, Almundher Al-Maawali, Anita H. Corbett

## Abstract

The RNA exosome is an evolutionarily conserved complex required for both precise RNA processing and decay. Mutations in *EXOSC* genes encoding structural subunits of the complex are linked to several autosomal recessive disorders. Here, we describe a missense allele of the *EXOSC4* gene, which causes a collection of clinical features in two affected siblings. This missense mutation (NM_019037.3: exon3:c.560T>C), changes a leucine residue within a highly conserved region of EXOSC4 to proline (p.Leu187Pro). The two affected individuals presented with prenatal growth restriction, failure to thrive, global developmental delay, intracerebral and basal ganglia calcifications, and kidney failure. Homozygosity for the damaging variant was identified through exome sequencing and Sanger sequencing confirmed segregation. To explore the functional consequences of this amino acid change, we modeled EXOSC4-L187P in the corresponding budding yeast protein, Rrp41 (Rrp41-L187P). Cells that express Rrp41-L187P as the sole copy of the essential Rrp41 protein show significant growth defects. The steady-state level of both the Rrp41-L187P and the EXOSC4-L187P proteins is significantly decreased compared to control Rrp41/EXOSC4. Consistent with this observation, targets of the RNA exosome accumulate in *rrp41-L187P* cells, including the 7S precursor of 5.8S rRNA. Polysome profiles show a significant decrease in translation in *rrp41-L187P* cells as compared to control cells with apparent incorporation of 7S pre-rRNA into polysomes. Taken together, this work adds the EXOSC4 subunit of the RNA exosome to the structural subunits of this complex that have been linked to human disease and defines foundational molecular defects that could contribute to the adverse growth phenotypes caused by this novel EXOSC4 pathogenic variant.

## Introduction

The RNA exosome is an evolutionarily conserved, ubiquitously expressed complex (1) composed of nine structural subunits and a catalytic 3’-5’ exonuclease/endonuclease (2, 3), which is required for essential processes such as the production of mature rRNA in the nucleus and turnover of various RNAs in both the cytoplasm and the nucleus (4). The nine structural subunits of the complex consist of three cap subunits (EXOSC1-3), and six core subunits (EXOSC4-9), which form a barrel-like structure (5). Recently, a number of pathogenic missense mutations have been identified in genes that encode these structural subunits of the RNA exosome complex. The initial finding was that mutations within the *EXOSC3* gene, which encodes one of the cap subunits, are linked to pontocerebellar hypoplasia type 1B (PCH1b) (6). Subsequent studies have led to the identification of a number of additional pathogenic mutations in *EXOSC3* (7) as well as mutations in *EXOSC1*, *2*, *5*, *8*, and *9* that have been linked to autosomal recessive disease (8-13). The majority of the pathogenic mutations identified in *EXOSC* genes cause single amino acid substitutions. The clinical presentation associated with these different mutations are diverse (14); however, many of the mutations impact the cerebellum at least to some extent, consistent with the link to pontocerebellar hypoplasia. How mutations that impact one essential complex cause distinct pathology is not yet clear.

Here, we report the identification of a missense mutation, p.L187P, in the *EXOSC4* gene that is linked to a neurodevelopmental disorder in two siblings. To explore the functional consequences of this amino acid substitution, we model this pathogenic mutation in the corresponding budding yeast protein Rrp41. Cells that express Rrp41-L187P as the sole copy of the essential Rrp41 protein show significant growth defects. The steady-state level of the Rrp41-L187P protein is decreased relative to the level of wildtype Rrp41, suggesting the L187P amino acid change could alter protein stability. Consistent with these results, *rrp41-L187P* cells show significant accumulation of RNAs that are normally processed or degraded by the RNA exosome, including the 7S precursor of mature 5.8S rRNA as well as reduced overall translation. Finally, steady-state levels of the mammalian EXOSC4-L187P protein are decreased relative to wildtype EXOSC4 in a neuronal cell line, suggesting that mechanisms defined in budding yeast may extend to the human protein. This study adds EXOSC4 to the RNA exosome subunits that have now been linked to neurological deficits in humans and defines molecular consequences from this disease-causing EXOSC variant.

## Results

### Clinical report

The pedigree for a family with two affected siblings born to consanguineous parents is depicted in **Figure 1**. The index patient (III-1; **Figure 1A**) is a female; she was the first child born to a 20-25-year-old primigravida mother, following a pregnancy complicated with oligohydramnions and intra-uterine growth restriction. She was delivered via a planned Caesarean section at 36 weeks of gestation, with a birth weight of 2.06 kg (10th percentile), head circumference of 31.8 cm (25th percentile), and length of 41 cm (3rd percentile). She required no active resuscitation and had normal APAGR scores. At initial evaluation, the patient presented with developmental delay and esotropia. Her developmental milestones assessment showed a global delay in all domains with progressive postnatal growth retardation. Her physical examination revealed frontal bossing, deep-seated eyes, retrognathia and loss of facial subcutaneous fat as well as sparse hair (data not shown). A neurological examination showed axial hypotonia, all limbs spasticity with brisk deep tendon reflexes but absent clonus. Her systemic examinations including the abdomen, heart and skin were unremarkable. Ophthalmology examination showed large angle esotropia with both eyes in the adduction position and no significant refractive error.

**Figure 1:**
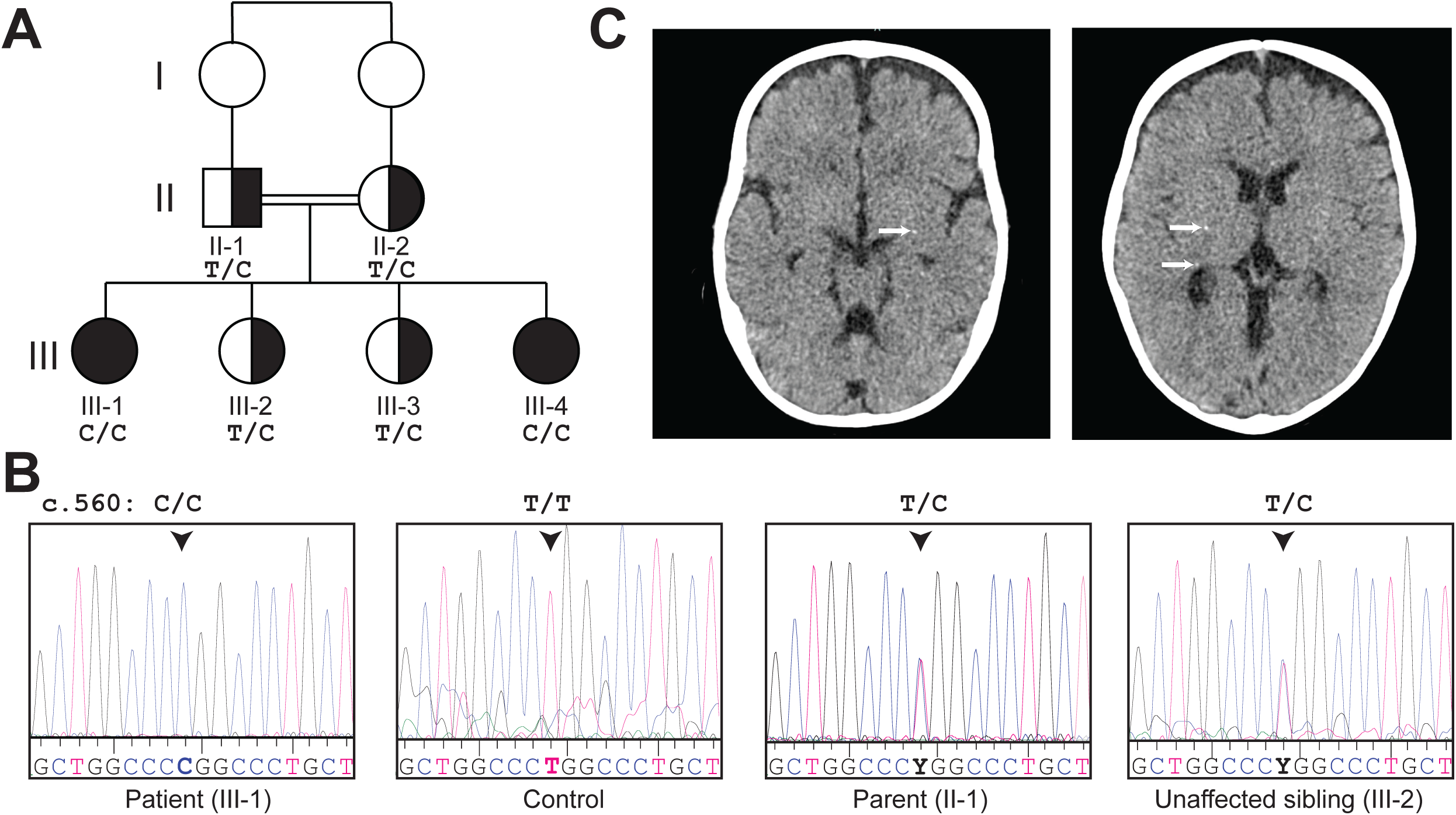
A novel EXOSC4 variant. (A) Pedigree of the family with two affected offspring (III-1 and III-4) and two unaffected siblings (III-2 and III-3). Fully shaded symbols indicate affected individuals. The genotype is shown for those individuals who were tested. All genotypic carrier individuals are phenotypically healthy. (B) Chromatograms for the Sanger sequencing showing the sequence variants present in Patient, Control samples, Parent, and Unaffected sibling. The sequencing identified the *EXOSC4* variant (NM_019037.3:c.560T>C (p.Leu187Pro). (C) Axial CT brain scan for the affected individual III-4 at the age of 13 months. A few tiny foci of hyperattenuation are seen in both parietal lobes and in basal ganglia bilaterally (Arrows). There is prominence of extracerebral CSF spaces in the frontoparietal region bilaterally indicating mild volume loss and brain atrophy.

At the age of 10-15 years, the development milestones for patient III-1 showed ability to walk with support only, although she could stand alone. She was able to hold pencils and scribble, but she could not draw or copy a circle. She was able to say two-word sentences and understand simple commands. She could count up to three and identify body parts and a few colors. Her overall developmental stage was at ∼12-24 months. Her routine examinations revealed iron deficiency anemia, otherwise normal liver functions, bone profile, serum calcium and magnesium, thyroid function tests, karyotype, metabolic screen, urine organic acid and other routine tests. Echocardiography, electrocardiogram, X-ray skeletal survey, nerve conduction studies, and ultrasound abdomen and pelvis and hearing test were all normal. A CT brain imaging showed bilateral diffuse calcification of the basal ganglia including the caudate nucleus along with subcortical calcification of both parietal lobes at the centrum semiovale level. No other brain malformations were identified and the ventricular system and posterior structures fossa appeared normal. The MRI brain showed mild generalized brain atrophy with no other abnormalities (data not shown).

While this patient was under follow-up, a similarly affected sister (III-4; **Figure 1A**) was delivered to this consanguineous couple. She presented with a similar phenotype. Her pregnancy and delivery history were unremarkable and she was born at full term with normal birth parameters. At the first assessment, her growth parameters showed a weight of 9.3 kg (-2.5 SD), a head circumference of 47 cm (-0.33 SD), and a length of 80.5 cm (- 1.5 SD). Her physical examination revealed deep-seated eyes and retrognathia with pointed chin. Her neurological examination showed axial hypotonia, all limbs spasticity with brisk deep tendon reflexes but absent clonus. Her systemic examinations including the abdomen, heart and skin were unremarkable. Ophthalmology examination showed bilateral esotropia and myopic astigmatism. At follow-up, her growth metrics were a weight of 15.4kg (-3.7 SD), a head circumference of 49 cm (-2.5 SD), and a length of 107 cm (-4.8 SD). At a developmental assessment at the age of 10-15 years, she was unable to walk with assistance with an unsteady gait and she had frequent falls. She could go up and down the stairs with support only. She could hold a pencil and scribble and copy circle. She expressed her needs in 2–3-word sentences but with limited vocabulary. She responded to her name and could count from 1-3 only and identify a few colors. All investigations similar to the sister were within normal limits. However, the CT scan of the brain was reported as a few tiny foci of hyperattenuation detected in both parietal lobes and in basal ganglia bilaterally (**Figure 1B**).

### Whole exome analysis

A targeted genetic investigation of the initial patient included a Next Generation Sequencing (NGS) Panel analyzing genes linked to DNA repair (gene sequencing with deletion and duplication analysis of *DDB1, DDB2, ERCC1, ERCC2, ERCC3, ERCC4, ERCC5, ERCC6, ERCC8, GTF2H5, LIG4, MPLKIP, NHEJ1, POLH, UVSSA,XPA, XPC, XRCC4*), but no change was detected in any of these genes. Thus, exome sequencing was performed to identify the genetic basis for this condition. The exome sequencing analysis of the two affected subjects along with the normal parent revealed homozygosity for a variant in the *EXOSC4* gene that was confirmed by Sanger sequencing and co-segregate with the phenotype (**Figure 1C**). The candidate variant (NM_019037.3:c.560T>C (p.Leu187Pro) was the only variant shared between the two affected subjects and met the filtration criteria. Additionally, other genes known to cause neurodevelopmental delay, brain calcifications or failure to thrive were analyzed and no other pathogenic variants were identified. This variant is absent from the publicly available variant databases (1000 Genomes, Exome Variant Server, and gnomAD) and the in-house control database. This variant is predicted to be damaging/pathogenic by multiple in-silico prediction tools include BayesDel, MetaRNN, BayesDel noAF, REVEL GERP, LRT, MutationAssessor, MutationTaster, SIFT, and PROVEAN. This site is highly conserved among 100 vertebrate genomes (including humans).

### Functional consequences of the L187P pathogenic amino acid change

As shown in **Figure 2A**, Leu 187 is located in a highly conserved region of the EXOSC4 protein. Within the three-dimensional structure of the RNA exosome complex, Leu187 lies in a region of EXOSC4 that is in close proximity to the interface with EXOSC9 (**Figure 2B**). Specifically, Leu187 of EXOSC4, which is located in an α-helix, interacts with Leu199 within a neighboring β-strand that is in contact with EXOSC9-I234. Similar types of contacts are formed in the yeast protein, Rrp41 in which L187 interacts with Rrp41-L199 that interacts with Rrp45-V248. Modeling of the L187P using the available human RNA exosome structure (PDB ID: 6D6R) (15) with Alphafold (16) does not predict major structural changes from wildtype EXOSC4 but predicts disruption of the specific above mentioned interactions which could result in destabilization of EXOSC4 and affect interaction with EXOSC9.

**Figure 2:**
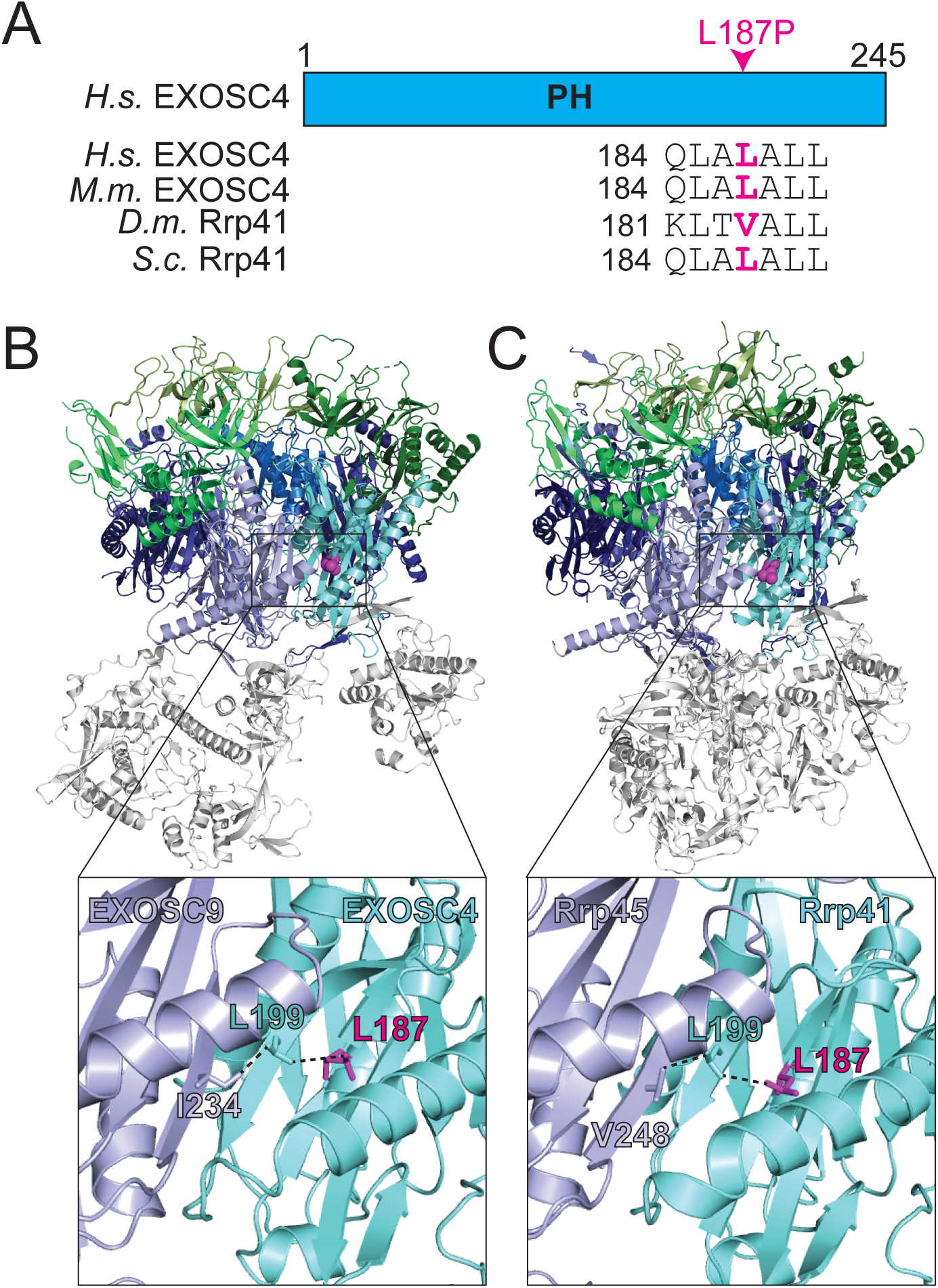
Conservation and location of the L187 residue in the EXOSC4/Rrp41 subunit of the RNA exosome that is altered in disease. (A) Schematic of domain organization of EXOSC4. EXOSC4 is composed of a single PH-like domain. Alignment of EXOSC4/Rrp41 amino acid sequences from *Homo sapiens* (*H.s.*), *Mus musculus* (*M.m.*), *Drosophila melanogaster* (*D.m.*), and *Saccharomyces cerevisiae* (*S.c.*) below the schematic show the conservation of the L187 residue (pink/magenta). (B) Structural model of the human RNA exosome complex [PDB: 6D6R; (15)] and (C) structural model of the yeast RNA exosome complex [PDB: 6FSZ; (17)]. The zoomed in boxes in (B) and (C) depict the exosome ring subunits EXOSC4/Rrp41 (Teal) and EXOSC9/Rrp45 (Purple) that share an interface and shows the location of the L187 residue (pink/magenta) in EXOSC4/Rrp41. The EXOSC4/Rrp41 residue L199 (Teal) and the EXOSC9 residue I234/Rrp45 residue V248 (Purple) in close contact with the EXOSC4/Rrp41 residue L187 are shown.

To explore the functional consequences of the L187P amino acid substitution in the EXOSC4 subunit of the RNA exosome, we modeled this pathogenic variant in the budding yeast orthologue of EXOSC4, Rrp41. As shown in **Figure 2A**, L187 is conserved between human EXOSC4 and budding yeast Rrp41. Furthermore, the conserved structure of the RNA exosome complex (17) means that Rrp41 L187 lies in close proximity to the interface with the core subunit that corresponds to EXOSC9, Rrp45 (**Figure 2C**). The *rrp41-L187P* budding yeast cells were produced using either a plasmid shuffle or CRISPR genome editing approach to express this Rrp41 variant as the sole copy of the essential Rrp41 protein. As an initial test of the function of Rrp41-L187P, we examined cell growth of these *rrp41-L187P* cells. Results of this analysis reveal that *rrp41-L187P* cells show growth defects at all temperatures examined with virtually no growth at 37°C, severely impaired growth at 30°C, and a mild growth defect even at 25°C as compared to control *RRP41* cells (**Figure 3A**). To examine whether the L187P amino acid substitution alters the Rrp41 protein level, both wildtype Rrp41 and Rrp41-L187P were Myc-tagged and analyzed by immunoblotting. As shown in **Figure 3B** and quantitated in **Figure 3C**, Rrp41-L187P levels are decreased by more than 50% compared to Rrp41 at both 30°C and 37°C. To assess the function of the RNA exosome in the *rrp41-L187P* cells, we examined the steady-state levels of documented RNA exosome transcript targets (18-20). For three different RNA targets analyzed, there is significant accumulation (10-20-fold increase) of the RNA analyzed in *rrp41-L187P* cells as compared to *RRP41* cells (**Figure 3D-F**), suggesting that RNA exosome function is impaired in these cells.

**Figure 3:**
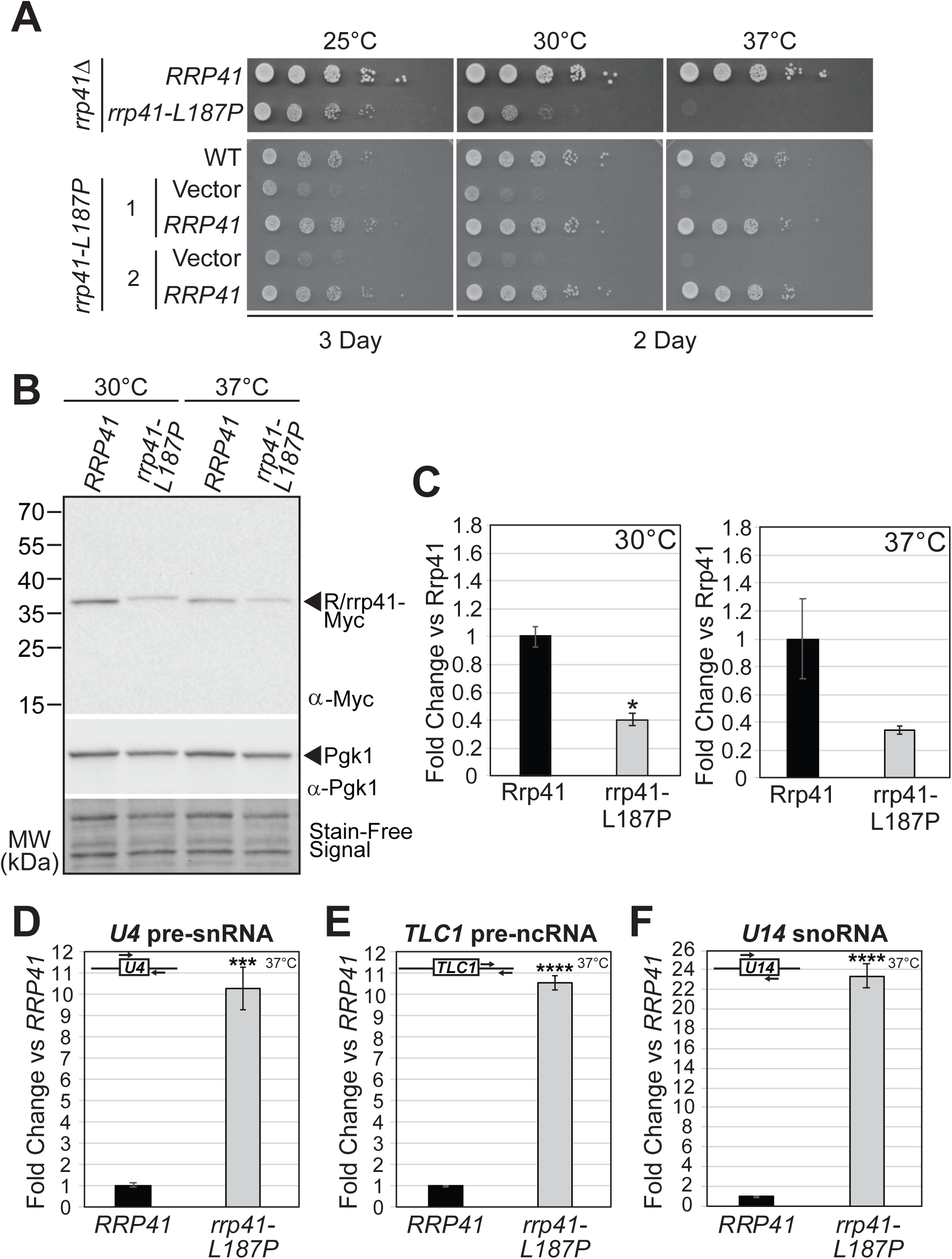
The budding yeast rrp41-L187P variant modeling the EXOSC4-L187P variant identified in patients causes impaired growth, reduced Rrp41 protein level, and elevated levels of RNA exosome target transcripts in *S. cerevisiae* at 37°C. (A) The *rrp41-L187P* mutant cells show impaired growth at all temperatures (25, 30, 37°C) tested. In the Upper Panel, in a solid media growth assay, *S. cerevisiae rrp41Δ* mutant cells expressing *rrp41-L187P* variant as the sole copy of *RRP41* show impaired growth compared to cells expressing *RRP41* wildtype control at all temperatures tested. The *rrp41Δ* cells containing *RRP41* or *rrp41-L187P* plasmid were serially diluted, spotted onto minimal media, and grown at indicated temperatures for 2-3 days. In the Lower Panel, in a solid media growth assay, two CRISPR/Cas9-engineered *S. cerevisiae rrp41-L187P* mutant strains show impaired growth compared to wildtype cells at all temperatures tested and their growth can be rescued by wildtype *RRP41* expression from a plasmid. Two *rrp41-L187P* CRISPR mutant strains (1 & 2) containing Vector or *RRP41* plasmid and wildtype (WT) cells were serially diluted, spotted onto minimal media, and grown at indicated temperatures for 2-3 days. Construction of the *rrp41-L187P* CRISPR mutant strains is described in Materials and Methods. (B) The steady-state level of the rrp41-L187P protein variant is significantly decreased at 30°C and 37°C. Lysates of *rrp41Δ* cells solely expressing Myc-tagged, wild-type Rrp41 or rrp41-L187P grown at 30°C or 37°C were analyzed by immunoblotting with an anti-Myc antibody to detect Rrp41-Myc and an anti-Pgk1 antibody to detect 3-phosphoglycerate kinase (Pgk1) as a loading control. The Stain-Free signal on the immunoblot is also shown as a loading control. (C) Quantitation of the fold change in rrp41-L187P protein level relative to Rrp41 detected in lysates of *rrp41Δ* cells solely expressing Myc-tagged Rrp41 or rrp41-L187P grown at 30°C or 37°C from two immunoblot experiments – one represented in (B). Graph shows the mean fold change of Rrp41-Myc protein level relative to wildtype Rrp41 from two independent experiments (n = 2). Error bars represent standard error of mean. Statistical significance is denoted by asterisk (*P-value ≤ 0.05). (D) The *rrp41-L187P* cells show elevated, steady-state levels of *U4* pre-snRNA, *TLC1* telomerase component pre-RNA, and *U14* snoRNA relative to *RRP41* cells at 37°C. Total RNA was isolated from *rrp41Δ* cells solely expressing *RRP41* or *rrp41-L187P* grown at 37°C and transcript levels were measured by RT-qPCR using gene specific primers, normalized relative to *RRP41*, and graphed as described in Materials and Methods. Gene specific primer sequences are summarized in Table 2 and their locations within the transcript are graphically represented by the cartoons in the top left of each bar graph. Within each cartoon transcript, the box represents the body of the mature transcript. Error bars represent standard error of the mean from three biological replicates. Statistical significance of the RNA levels in *rrp41-L187P* cells relative to *RRP41* cells is denoted by an asterisk (***P-value ≤ 0.001; ****P-value ≤ 0.0001).

### rrp41-L187P cells show defects in rRNA processing and translation

To reveal the extent to which rRNA processing is impacted in *rrp41-L187P* cells, we performed northern blotting and used a series of probes (P1-P6) to analyze steps in rRNA processing (**Figure 4A**). These data reveal significant early rRNA processing defects resulting in accumulation of the 35S pre-rRNA transcript with a concomitant decrease in the levels of precursors of 18S and 25S, and accumulation of 7S pre-rRNA, the 5.8S precursor that is directly processed by the RNA exosome (21, 22). We therefore assessed how these significant ribosome biogenesis changes impact the translating ribosomes by performing polysome profiling (**Figure 4B**). Corroborating these findings, analyses of the polysome profiles reveal a marked reduction of polysome fractions. Further northern blot analysis of the polysome fractions indicates a significant amount of 7S rRNA containing particles in polysome fractions of the *rrp41-L187P* mutant cells (**Figure 4C**). To assess whether these 7S pre-rRNA containing complexes migrating in polysome fractions are aggregated complexes or premature subunits that escaped into the translating ribosome pool, we removed cycloheximide and added puromycin to lysates prior to analysis on gradients. Removal of cycloheximide from polysome analysis allows run off of the translating ribosomes and puromycin dissociates the ribosomes into the small and large subunits (23). Under these conditions, we observe that the bulk of 7S pre-rRNA and 5.8S rRNA shifts to the 80S and 60S fractions (**Figure 4D**), demonstrating that a significant portion of 7S pre-rRNA found in the polysome fractions of the *rrp41-L187P* mutant cells is associated with ribosomes.

**Figure 4:**
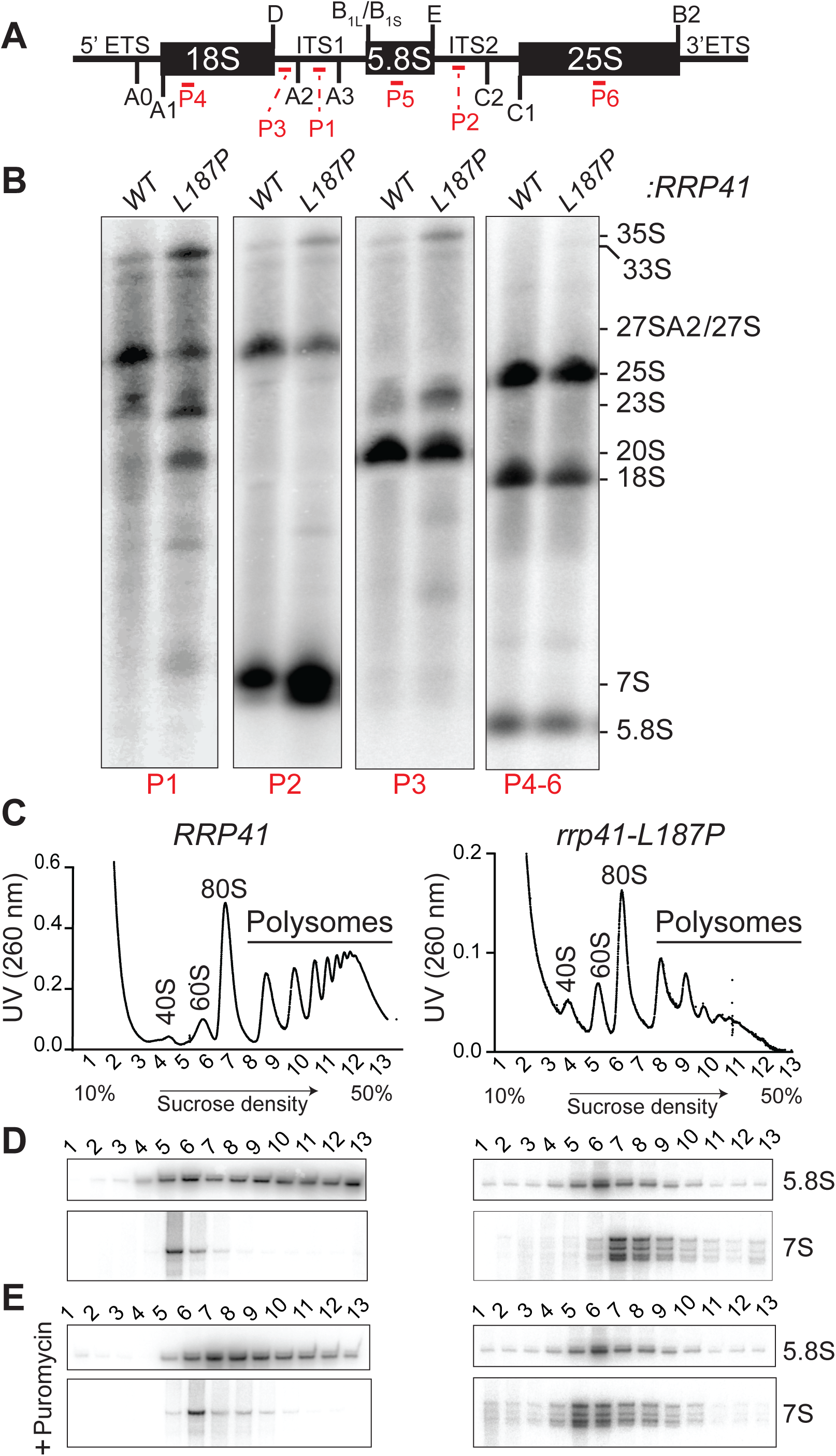
The *rrp41-L187P* mutant cells exhibit defects in rRNA processing and translation (A) Schematic of yeast 35S rRNA that highlights the locations of rRNA cleavage sites (A-E; Black) and northern blot probes (P1-P6; Red). The 35S rRNA transcript contains 18S, 5.8S, and 25S rRNA separated by internal transcribed spacer 1 and 2 (ITS1, ITS2) and flanked by 5’ and 3’ external transcribed spacer (5’ETS, 3’ETS). (B) Northern blotting reveals that *rrp41-L187P* mutant cells show defects in rRNA processing, including accumulation of 7S pre-rRNA. A northern blot on total RNA from *rrp41Δ* cells solely expressing wildtype Rrp41 (WT) or Rrp41-L187P variant (L187P) grown at 30°C was performed and probed with rRNA oligonucleotide probes P1, P2, P3, and P4-6 (Red). (C) Polysome profiling reveals that *rrp41-L187P* CRISPR mutant cells exhibit defects in translation at 37°C. Protein lysates of *rrp4-L187P* CRISPR mutant cells and *RRP41* wildtype cells were examined on sucrose density gradients. Clarified cell extracts were resolved on a 10-50% sucrose gradient and scanned at 260 nm. Peaks corresponding to the small (40S) and large (60S) ribosome subunits, as well as the monosomes (80S) and polysomes are marked. (D) Northern blotting on fractions from sucrose gradients treated with cycloheximide and (E) fractions from gradients not treated with cycloheximide and treated with puromycin reveal the distribution of 5.8S rRNA and 7S pre-rRNA.

### Steady-state levels of mammalian EXOSC4-L187P are reduced compared to wildtype EXOSC4

Our results modeling the novel disease-associated EXOSC4-L187P in budding yeast show that the L187P amino acid substitution in Rrp41 decreases the steady-state level of the protein as shown in **Figure 3B,C**. To assess whether this amino acid change alters the steady-state levels of the mammalian EXOSC4 protein, we expressed Myc-tagged wild-type mouse EXOSC4 or EXOSC4-L187P in cultured Neuro2A cells and examined the levels of protein by immunoblotting (**Figure 5A**). As shown in the quantitation presented in **Figure 5B**, the EXOSC4-L187P level is ∼70% of the control EXOSC4 protein level, suggesting that the L187P amino acid substitution could affect the stability of the EXOSC4 protein, albeit not to the same extent observed for yeast Rrp41.

**Figure 5:**
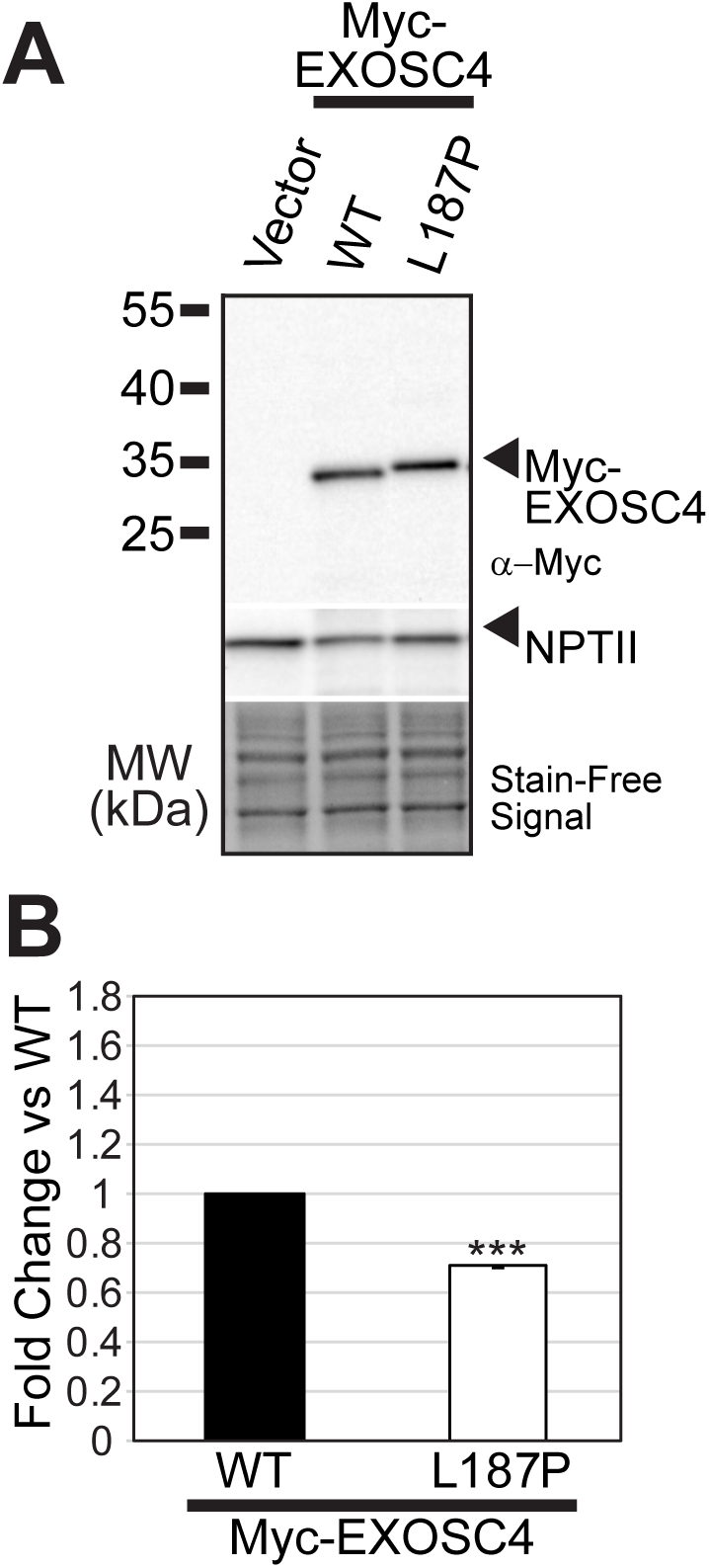
(A) The murine EXOSC4-L187P variant, corresponding to the human EXOSC4 variant identified in patients, is expressed at a lower level compared to wild-type murine EXOSC4 in a mouse neuronal cell line. The steady-state level of Myc-tagged murine EXOSC4-L187P variant is lower relative to Myc-tagged wild-type EXOSC4 (WT) in mouse N2a cells. Lysates of mouse N2a cells transfected with empty vector or vector expressing murine Myc-EXOSC4 or Myc-EXOSC4-L187P were analyzed by immunoblotting with anti-Myc antibody to detect Myc-EXOSC4 proteins. The Stain Free signal serves as a loading control and neomycin phosphotransferase II (NPTII) serves as a transfection control. (B) Quantitation of the fold change in EXOSC4-L187P protein level relative to EXOSC4 detected in lysates of N2a cells expressing Myc-tagged EXOSC4 or EXOSC4-L187P from two immunoblot experiments – one represented in (A). Graph shows the mean fold change of EXOSC4-Myc protein level relative to wildtype EXOSC4 (WT) from two independent experiments (n = 2). Error bars represent standard error of mean. Statistical significance is denoted by asterisk (***P-value ≤ 0.001).

## Discussion

Here, we report for the first time a pathogenic mutation in *EXOSC4*, which encodes a structural subunit of the RNA exosome. This work adds the *EXOSC4* gene to a growing list of genes that are linked clinically to a class of syndromes termed exosomopathies. Consistent with many of the exosomopathies reported thus far (8, 14), the pathogenic mutation identified in *EXOSC4* (p.L187P) causes a single amino acid change in an evolutionary conserved sequence within EXOSC4. Modeling of this pathogenic amino acid substitution in the budding yeast protein corresponding to EXOSC4, Rrp41, reveals that cells which express the Rrp41-L187P variant as the sole copy of the essential Rrp41 protein show severe growth defects as well as accumulation of a number of RNA exosome target transcripts, including 7S pre-rRNA. The *rrp41-L187P* cells also show defects in translation and incorporation of 7S pre-rRNA into polysomes, suggesting a potential mechanism contributing to pathology in cells with defective RNA exosome function.

The two individuals described in this study show a number of clinical consequences that are not present in siblings that are heterozygous for the pathogenic mutation. Consistent with a global requirement for the RNA exosome to support key cellular functions such as maturation of rRNA to produce ribosomes, these individuals show defects in a number of different organs as well as failure to thrive. A number of exosomopathies have been associated with cerebellar defects. Why mutations that impair RNA exosome function would cause tissue- or cell-specific consequences is not yet clear as the requirement for RNA exosome function in different cell types and developmental programs is only starting to be defined (24-26). Some initial studies suggest that the function of the RNA exosome is critical to ensure proper differentiation for a number of different cell types ranging from B cells to blood cells (26, 27).

As in many clinical reports associated with mutations in RNA exosome genes (14), a single amino acid change within a core structural subunit of the RNA exosome causes pathology in the case reported here. Such single amino acid changes can alter protein function through multiple mechanisms. While the L187P change in EXOSC4/Rrp41 is not predicted to alter the overall structure of either the individual protein or the RNA exosome complex based on modeling in Alphafold, experimental evidence from both budding yeast (**Figure 3B,C**) and cultured mammalian cells (**Figure 5A,B**) shows that the steady state level of Rrp41-L187P/EXOSC4-L187P is decreased relative to control Rrp41/EXOSC4 protein. These results suggest that this amino acid change could alter the stability of the subunit or the overall complex. As EXOSC4/Rrp41 L187 lies within close proximity to the interface with another subunit of the core, EXOSC9/Rrp45, the change to proline could disrupt this interaction and destabilize the complex. Further studies will be required to probe the integrity of the RNA exosome complex in cells that express this EXOSC4/Rrp41 variant. Several other studies that have analyzed steady-state levels of RNA exosome subunits in patient-derived cells show decreased levels of not only the RNA exosome subunit effected by the disease mutation but also other subunits (13). Given the differences in the clinical presentation of individuals diagnosed with exosomopathies, a simple explanation that all result from a drop in the activity of the RNA exosome due to loss of the complex or individual subunits seems not sufficient. Likely the pathological consequences result from some combination of a decrease in the pool of active RNA exosome complex and specific consequences due to the pathologic amino acid change.

This study extends the analysis of RNA exosome target transcripts to explore the consequences for translation. Maturation of the 3′ end the 5.8S rRNA from the 7S pre-rRNAs involves a series of cleavage steps by different nucleases (28). The trimming of 7S to 6S pre-rRNA happens in the nucleus whereas the final processing of 6S pre-rRNA to 5.8S rRNA is cytoplasmic (22). As 3’-end trimming of the 7S pre-rRNA to the mature 5.8S required to assemble functional ribosomes by the RNA exosome is one of the best defined function of this complex, defects in this process would be predicted to alter translation. Indeed, a recent study demonstrated that several different budding yeast models of exosomopathies impair translation albeit, there appear to be differences in how translation is affected in different yeast models of exosomopathies (29). The analysis of the core subunit here is consistent with the previous finding (29), suggesting that mutations that affect the core subunits vs the cap subunits of the RNA exosome cause distinct translational defects with defects in core subunits such as Rrp41 causing an overall reduction of ribosomes while defects in cap subunits leading to formation of halfmers indicating a potential problem in 60S maturation or subunit joining. Further studies will be required to determine whether these changes in translation are confirmed in patient cells or mammalian disease models as well as how these molecular changes alter the proteome. Ultimately, the goal is to understand how fundamental changes in RNA exosome function and the downstream consequences of these changes are linked to clinical consequences associated with exosomopathies.

In summary, the work presented her expands the group of disorders termed exosomopathies by reporting a neurodevelopmental disorder in two sisters homozygous for a pathogenic missense mutation in the *EXOSC4* gene. Studies exploring the consequences of this pathogenic amino acid substitution (L187P) in a budding yeast model reveal defects in the function of the RNA exosome, including dysregulation of translation. A major challenge remains defining how an essential complex that contributes to critical biology in all cell types causes distinct pathological consequences.

## Experimental Procedures

### Human Subjects

This study was approved by the Medical Research Ethical Committee of the Sultan Qaboos University (SQU MREC#1362). Parents provided written informed consent to participate and to publish their family pedigrees and clinical data. Clinical investigations were conducted according to the principles expressed in the Declaration of Helsinki. Genetic analyses were performed in accordance with bioethics rules of national laws.

### Exome sequencing

We performed whole-exome sequencing (WES) analysis on DNA isolated from blood from the two affected siblings and the father as a trio-exome analysis. DNA was barcoded and enriched for the coding exons of targeted genes using hybrid capture technology (Agilent SureSelect Human All-exons-V6). Prepared DNA libraries were then sequenced using Next-Generation Sequencing (NGS) technology [NovaSeq6000, 150 bp paired-end, at 200X coverage]. The reads were mapped against UCSC GRCh37/hg19 by Burrows-Wheeler Aligner (BWA 0.7.12). Genome Analysis Tool Kit (GATK 3.4) was used for variant calling. Variant filtration was applied to keep novel or rare variants (≤ 1%). Publicly available variant databases (1000 Genomes, Exome Variant Server, and GnomAD) and an in-house database of 1564 exomes were used to filter out common or benign variants specific to the Omani population. Only coding or splicing variants were considered. The phenotype and mode of inheritance (autosomal recessive) were considered. Variants of high impact or highly damaging missense, a CADD score ≥ 20 and shared between the affected individuals were prioritized. Sanger sequencing was used to confirm segregation. The *EXOSC4* forward and reverse primer sequences (AM1 & AM2) used for amplification of the target on *EXOSC4* are listed in Table 2.

### Chemicals and media

All chemicals were obtained from Sigma-Aldrich (St. Louis, MO), United States Biological (Swampscott, MA), or Fisher Scientific (Pittsburgh, PA) unless otherwise noted. All media were prepared by standard procedures (30).

### Protein structure analysis

We used the cryo-EM structure (PDB 6h25) of the human nuclear RNA exosome at 3.8Å resolution (31). Structural modeling was performed using the PyMOL viewer (The PyMOL Molecular Graphics System, Version 2.0 Schrödinger, LLC.). The mCSM webserver was used for predicting the effect of missense mutation (32).

### S. cerevisiae strains and plasmids

All DNA manipulations were performed according to standard procedures (33). *S. cerevisiae* strains and plasmids used in this study are listed in Table 1. The haploid *rrp41Δ* yeast strain (yAV2493) was generated by transformation of a heterozygous diploid *RRP41/rrp41Δ* yeast strain (BY4743 strain background; Open Biosystems, Huntsville, AL) with a wild-type *RRP41 URA3* maintenance plasmid (*CEN6*) and sporulation of the diploid to yield *rrp41Δ* haploid progeny containing the *RRP41* maintenance plasmid. The wild-type *RRP41 LEU2 CEN6* plasmid (pAC4179) and *RRP41 LEU2* 2µ plasmid (pAC2971) were generated by PCR amplification of the *RRP41* gene containing its endogenous promoter, 5’UTR, and 3’UTR from yeast genomic DNA using yeast gene specific primers (Integrated DNA Technologies) and cloning into pRS315 (34) and pRS425 (35), respectively. The *RRP41-Myc LEU2* plasmid (pAC4242; *CEN6*) was generated by PCR amplification of the *RRP41* promoter/coding sequence and *2xMyc*-*RRP41* 3’UTR products using the *RRP41* plasmid (pAC4179) and yeast gene specific primers, one of which encoded the 2xMyc tag, and cloning into pRS315 (34). The *rrp41-L187P LEU2* (pAC4180) and *rrp41-L187P-Myc* (pAC4243) variant *LEU2* plasmids were generated by site-directed mutagenesis of *RRP41 LEU2* (pAC4179) and *RRP41-Myc LEU2* (pAC4243) plasmid, respectively, using oligonucleotides encoding the L187P amino acid change (AC9500 & AC9501; Integrated DNA Technologies) and QuickChange II SDM Kit (Agilent). The pcDNA3-Myc-*Exosc4* (pAC3517) plasmid was generated by PCR amplification of the mouse *Exosc4* coding sequence from Neuro-2a cDNA using gene specific primers (Integrated DNA technologies) and cloning into pcDNA3 (Invitrogen) plasmid containing pCMV promoter and N-terminal Myc tag. The pcDNA3-Myc-*Exosc4-L187P* (pAC4231) variant plasmid was generated by site-directed mutagenesis of pcDNA3-Myc-*Exosc4* (pAC3517) plasmid using oligonucleotides encoding the L187P amino acid change (AC9505 & AC9506; Integrated DNA Technologies) and QuickChange II SDM Kit (Agilent). The pRS316-TEF1p-Cas9-CYC1t-SNR52p (pAC3846) pCas9 plasmid (*URA3*, *CEN6*), which derives from p414-*TEF1p-Cas9-CYC1t* plasmid [Addgene #43802; (36)] and p426-SNR52p-gRNA.CAN1.Y-SUP4t plasmid [Addgene #43803; (36)] has been described previously (29). The pRS316-*TEF1p-Cas9-CYC1t-SNR52p-RRP41_565.gRNA-SUP4t* (pAC4339) pCas9 plasmid containing the gRNA for targeting *RRP41* was constructed by PCR amplification of *RRP41_565.gRNA* with oligonucleotides AC9891 and AC6809 using p426-SNR52p-gRNA.CAN1.Y-SUP4t plasmid template (Addgene #43803) and cloning of SphI/KpnI-digested gRNA product into pAC3846 digested with SphI/KpnI. All plasmids were sequenced to ensure the presence of desired mutations and absence of any other mutations.

**Table 1.** Saccharomyces cerevisiae strains and plasmids.

| Strain/Plasmid | Description | Reference |
| --- | --- | --- |
| BY4741 (ACY402) | *MATa ura3Δ0 leu2Δ0 his3Δ1 met15Δ0* |  |
| *rrp41Δ* (yAV2493) | *MAT? ura3Δ0 leu2Δ0 his3Δ1 rrp41Δ::NEO [RRP41, URA3]* | This Study |
| *rrp41-L187P* (ACY3125) | *MATa ura3Δ0 leu2Δ0 his3Δ1 met15Δ0 rrp41-L187P* | This Study |
| *rrp41-L187P* (ACY3126) | *MATa ura3Δ0 leu2Δ0 his3Δ1 met15Δ0 rrp41-L187P* | This Study |
| pRS315 | *CEN6, LEU2, amp<sup>R</sup>* | (34) |
| pRS425 | *2μ, LEU2, amp<sup>R</sup>* | (35) |
| pAC2971 | *RRP41, LEU2, 2μ* | This Study |
| pAC4179 | *RRP41-Native 3'UTR in pRS315, CEN6, LEU2, amp<sup>R</sup>* | This Study |
| pAC4180 | *rrp41-L187P-Native 3'UTR in pRS315, CEN6, LEU2, amp<sup>R</sup>* | This Study |
| pAC4242 | *RRP41-2xMyc-Native 3'UTR in pRS315, CEN6, LEU2, amp<sup>R</sup>* | This Study |
| pAC4243 | *rrp41-L187P-2xMyc-Native 3'UTR in pRS315, CEN6, LEU2, amp<sup>R</sup>* | This Study |
| pcDNA3 | *pCMV, NeoR, amp<sup>R</sup>* | Invitrogen |
| pAC3517 | *Myc-Exosc4 in pcDNA3, NeoR, amp<sup>R</sup>* | This Study |
| pAC4231 | *Myc-Exosc4-L187P in pcDNA3, NeoR, amp<sup>R</sup>* | This Study |
| pAC3846 | *TEF1p-Cas9-CYC1t-SNR52p in pRS316, CEN6, URA3, amp<sup>R</sup>* | (29) |
| pAC4339 | *TEF1p-Cas9-CYC1t-SNR52p-RRP41_565.gRNA-SUP4t in pRS316, CEN6, URA3, amp<sup>R</sup>* | This Study |

#### Generation of integrated rrp41-L187P mutant strains using CRISPR-Cas9 genome editing

Two *rrp41-L187P* (ACY3125; ACY3126) mutant strains were generated using CRISPR/Cas9 editing with a single pCas9-gRNA expression plasmid and double-stranded homology-directed repair (HDR) oligonucleotides in a wild-type BY4741 strain essentially as described before (36). The single pCas9-gRNA plasmid on a pRS316 (*URA3, CEN6*) backbone is derived from p414-*TEF1p-Cas9-CYC1t* plasmid (Addgene #43802) and p426-SNR52p-gRNA.CAN1.Y-SUP4t plasmid (Addgene #43803). Constitutive expression of Cas9 is driven by the *TEF1* promoter and constitutive expression of the gRNA Is driven by the *SNR52* promoter. Specifically, 500 ng of pAC3846 (pCas9 without gRNA), pAC4339 (pCas9 + *RRP41* gRNA) +/- 1 nmol of double-stranded *rrp41-L187P* HDR oligonucleotide (AC9893/9894) and 50 μg salmon sperm DNA was transformed into wild-type BY4741 cells by standard Lithium Acetate (LiOAc) transformation protocol (37). HDR oligonucleotides are listed in **Table 2**. Cells were plated on Ura^-^ minimal media plates and incubated at 30°C for 2 days. Large colonies on plates with cells transformed pCas9-gRNA and HDR oligonucleotides were restreaked to new Ura^-^ minimal media plates and screened for the presence of *rrp41-L187P* mutations via Sanger sequencing of genomic *RRP41*.

**Table 2.**
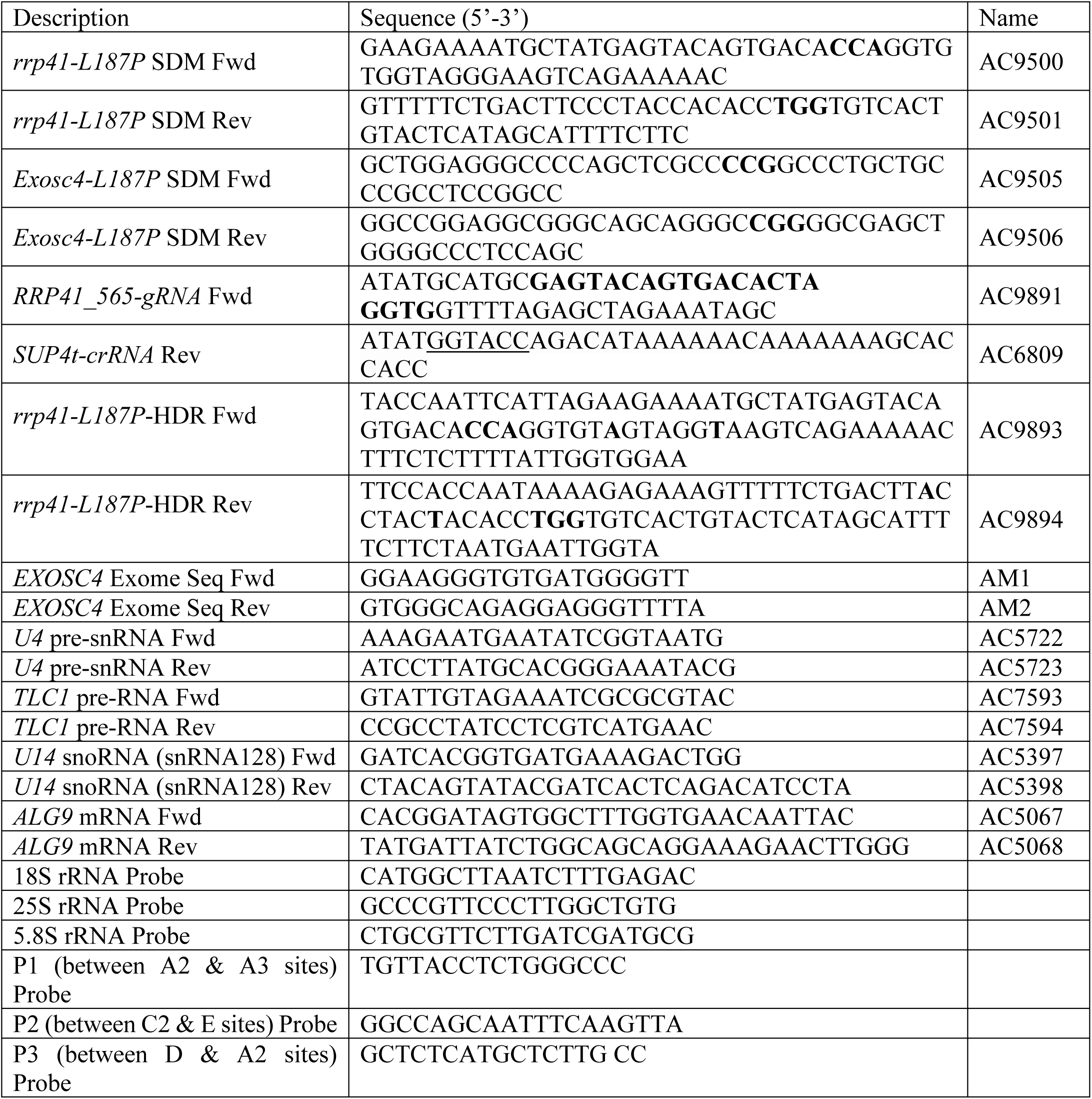
DNA oligonucleotide primers and probes.

### *S. cerevisiae* cell growth assays

The *rrp41Δ* yeast strain (yAV2493) was transformed with wild-type *RRP41* (pAC4179) or *rrp41-L187P* (pAC4180) variant *LEU2* plasmid and selected on Leu-plates. The Leu+ transformants were streaked and grown on a 5-flouroorotic acid (5-FOA) plate to select for cells that had lost the *RRP41 URA3* maintenance plasmid (38), and thus only contained the low copy, wild-type *RRP41 LEU2* plasmid or *rrp41-L187P LEU2* variant plasmid. The growth of the *rrp41Δ* cells containing *RRP41 LEU2* or *rrp41-L187P LEU2* plasmid was assayed on solid media by growth of cells in 2 mL Leu- minimal media overnight at 30°C, serial dilution of the cells (in 10-fold dilutions), spotting of cells on Leu- minimal media plates and incubation of the plates at 25°C, 30°C and 37°C for 2-3 days. The *rrp41-L187P* CRISPR mutant strains (ACY3125 & ACY3126) transformed with vector (pRS425) or *RRP41* (pAC2971) 2µ *LEU2* plasmid and wildtype (BY4741) cells transformed with vector (pRS425) were grown in 2 mL Leu- minimal media overnight at 30°C and similarly serially diluted, spotted onto Leu-plates, and incubated at 25°C, 30°C and 37°C for 2-3 days as indicated.

### Immunoblotting

For analysis of C-terminally Myc-tagged wild-type Rrp41 and rrp41-L187P variant protein expression levels, *rrp41* cells expressing control Rrp41-Myc (pAC4242) or rrp41-L187P-Myc (pAC4243) protein were grown in 2 ml Leu- minimal media overnight at 30°C to saturation and 10 ml cultures with an OD_600_ = 0.4 were prepared and grown at 30 or 37°C for 5 hr. Cell pellets were collected by centrifugation, transferred to 2 ml screw-cap tubes and stored at -80°C. For analysis of EXOSC4 expression levels, mouse N2a cells (39) were transiently transfected with pcDNA3 vector (Invitrogen) containing mouse wild-type *Myc-Exosc4* (pAC3517) or *Myc-Exosc4-L187P* variant (pAC4231) plasmid, or empty vector (pcDNA3; Invitrogen) using Lipofectamine 2000 (Invitrogen) and cells were collected 24 hr after transfection.

Budding yeast cell lysates were prepared by resuspension of cells in 0.5 ml RIPA-2 Buffer [50 mM Tris-HCl, pH 8; 150 mM NaCl; 0.5% sodium deoxycholate; 1% NP40; 0.1% SDS] supplemented with protease inhibitors [1 mM PMSF; Pierce™ Protease Inhibitors (Thermo Fisher Scientific)], addition of 300 µl glass beads, disruption in a Mini Bead Beater 16 Cell Disrupter (Biospec) for 4 x 1 min at 25°C, and centrifugation at 16,000 x *g* for 20 min at 4°C. Mouse N2a cell lysates were prepared by lysis in RIPA-2 Buffer and centrifugation at 16,000 x *g* for 10 min at 4°C. Protein lysate concentration was determined by Pierce BCA Protein Assay Kit (Life Technologies). Whole cell lysate protein samples (20-50 µg) were resolved on Criterion 4-20% gradient denaturing gels (Bio-Rad), transferred to nitrocellulose membranes (Bio-Rad) and Myc-tagged Rrp41 and EXOSC4 proteins were detected with anti-Myc monoclonal antibody 9B11 (1:2000; Cell Signaling). As loading controls, 3-Phosphoglycerate kinase (Pgk1) protein was detected with anti-Pgk1 monoclonal antibody (1:30,000; Invitrogen) and Stain-Free signal on the immunoblot was included. For transfection control, Neomycin phosphotransferase II (NPTII) expressed from *NeoR* cassette on pcDNA3*-Exosc4* vectors was detected with anti-NPTII monoclonal antibody (1:1000; Cell Applications, Inc.). Primary antibodies were detected using goat secondary antibodies coupled to horseradish peroxidase (1:3000; Jackson ImmunoResearch Inc) and enhanced chemiluminescence signals were captured on a ChemiDoc Imaging System (Bio-Rad).

### Quantitation of immunoblotting

The protein band intensities/areas from immunoblots were quantitated using ImageJ v1.4 software (National Institute of Health, MD; http://rsb.info.nih.gov/ij/) or ImageLab software (Bio-Rad) and mean fold changes in protein were calculated in Microsoft Excel for Mac 2011 (Microsoft Corporation). To quantitate and graph the mean fold change in rrp41-L187P-Myc variant level relative to wild-type Rrp41-Myc level in *rrp41Δ* cells grown at 30°C or 37°C from two immunoblots, Rrp41-Myc and rrp41-L187P-Myc intensity was first normalized to loading control Pgk1 intensity and then normalized to wildtype Rrp41-Myc intensity at 30°C or 37°C for each immunoblot. To calculate the mean fold change in Myc-EXOSC4-L187P variant relative to wildtype Myc-EXOSC4 in N2a cells from two immunoblots, Myc-EXOSC4-L187P and Myc-EXOSC4 intensity were first normalized to Stain Free signal and NPTII transfection control, and then normalized to wildtype Myc-EXOSC4 for each immunoblot. The mean fold change in rrp41-L187P-Myc level relative to Rrp41-Myc and Myc-EXOSC4-L187P level relative to Myc-EXOSC4, and standard error of the mean were calculated and graphically represented. The statistical significance of the fold change for rrp41-L187P-Myc variant relative to Rrp41-Myc at 30°C or 37°C and Myc-EXOSC4-L187P relative to Myc-EXOSC4 was calculated using the Student’s *t*-test on GraphPad software.

### Total RNA isolation from S. cerevisiae

To prepare *S. cerevisiae* total RNA from cell pellets of 10 ml cultures grown to OD_600_ = 0.5-0.8, cell pellets in 2 ml screw-cap tubes were resuspended in 1 ml TRIzol (Invitrogen), 300 µl glass beads were added, and cell samples were disrupted in Mini Bead Beater 16 Cell Disrupter (Biospec) for 2 min at 25°C. For each sample, 100 µl of 1-bromo-3-chloropropane (BCP) was added, the sample was vortexed for 15 sec, and incubated at 25°C for 2 min. Sample was centrifuged at 16,300 x *g* for 8 min at 4°C and upper layer was transferred to a fresh microfuge tube. RNA was precipitated with 500 µl isopropanol and sample was vortexed for 10 sec to mix. Total RNA was pelleted by centrifugation at 16,300 x *g* for 8 min at 4°C. RNA pellet was washed with 1 ml of 75% ethanol, centrifuged at 16,300 x *g* for 5 min at 4°C, and air dried for 15 min. Total RNA was resuspended in 50 µl diethylpyrocarbonate (DEPC (Sigma))-treated water and stored at -80°C.

### Quantitative RT-PCR

For analysis of *U4* pre-snRNA, *TLC1* pre-RNA, and *U14* snoRNA levels in wild-type *RRP41* and *rrp41-L187P* mutant cells, *rrp41Δ* (yAV2493) cells solely containing control *RRP41* (pAC4179) or *rrp41-L187P* (pAC4180) plasmid were grown in biological triplicate in 2 ml Leu- minimal media overnight at 30°C, 10 ml cultures with an OD_600_ = 0.4 were prepared and grown at 37°C for 5 hr. Cells were collected by centrifugation (2,163 x *g*; 16,000 x *g*), transferred to 2 ml screw cap tubes and stored at -80°C. Following total RNA isolation from each cell pellet, 1 µg RNA was reverse transcribed to 1^st^ strand cDNA using the M-MLV Reverse Transcriptase (Invitrogen) and 0.3 µg random hexamers according to manufacturer’s protocol. Quantitative PCR was performed on technical triplicates of cDNA (10 ng) from independent biological triplicates using gene specific primers to detect *U4* pre-snRNA, *TLC1* pre-RNA, and *U14* snoRNA (0.5 µM; Table 2) and QuantiTect SYBR Green PCR master mix (Qiagen) on a StepOnePlus Real-Time PCR machine (Applied Biosystems; T_anneal_ = 55°C; 44 cycles). Primers to detect *ALG9* mRNA were used as a normalization control (Table 2). The mean RNA levels were calculated by the ΔΔCt method (40), normalized to mean RNA levels in *RRP41* cells, and converted and graphed as RNA fold change relative to *RRP41* with error bars that represent the standard error of the mean.

### Northern blot analysis of rRNAs

For analysis of ribosomal RNAs, *rrp41Δ* (yAV2493) cells solely containing *RRP41* (pAC4179) or *rrp41-L187P* plasmid were grown to mid-log phase 30°C and total RNA was extracted using the hot phenol method (41). Northern blotting was carried out essentially as previously described (42) using the oligonucleotide probes – 18S, 25S, 5.8S, P1 (between the A2 and A3 sites), P2 (between the C2 and E sites), and P3 (between the D and A2 sites) – listed in Table 2.

### Polysome analysis

To analyze polysome profiles, wildtype (BY4741) and *rrp41-L187P* CRISPR mutant (ACY3125) cells were cultured to mid-log phase in YP-dextrose media at 37°C and harvested with or without the addition of 0.1 mg/ml cycloheximide. Subsequently, the cells were washed and lysed in an ice-cold gradient buffer composed of 20 mM HEPES, pH 7.4, 5 mM MgCl2, 100 mM NaCl, and 2 mM DTT, supplemented with complete protease inhibitor cocktail (Roche) and 0.1 mg/ml cycloheximide or no cycloheximide. Cryogenic grinding was used to break the cells, and the resulting cell lysate was cleared by centrifugation at 10,000 g for 10 minutes. The absorbance of the cleared lysate was measured at UV260, and an equal amount of lysate was applied to 10–50% sucrose gradients in gradient buffer for all samples. For samples lacking cycloheximide, 2.5 mM puromycin was added to the cleared cell lysate. The lysate was then incubated on ice for 15 minutes and subsequently placed at 37°C before being loaded onto the gradient. Gradients were centrifuged for 2 hours at 40,000 RPM in a SW41Ti rotor and fractionated using a BioComp fraction collector.

## Data Availability Statement

Strains and plasmids are available upon request. The authors affirm that all data necessary for confirming the conclusions of the article are present within the article, figures, and tables.

## ACKNOWLEDGEMENTS

We thank members of the research groups for project discussions and experimental insight. We appreciate the insightful conversations with Dr. Ambro van Hoof. This study was supported by NIH awards R35GM138123 to H.G, R01GM130147 to A.H.C and A.v.H. and a Synergy II Nexus Award provided by the Woodruff Health Sciences Center (WHSC), Emory School of Medicine, the Office of the Provost, and Emory College of Arts and Sciences (ECAS) to H.G. and A.H.C. L.A.C. was supported by NIH T32GM149422.

